# Long-Term Outcomes After Transcatheter Aortic Valve Replacement: Meta-Analysis of Kaplan-Meier-Derived Individual Patient Data

**DOI:** 10.1101/2023.03.20.23287505

**Authors:** Junichi Shimamura, Sho Takemoto, Shinichi Fukuhara, Yoshihisa Miyamoto, Yujiro Yokoyama, Hisato Takagi, Polydoros N Kampaktsis, Dhaval Kolte, Kendra J Grubb, Toshiki Kuno, Azeem Latib

## Abstract

**Background:** Transcatheter aortic valve replacement (TAVR) is as an alternative treatment to surgical AVR, but the long-term outcomes of TAVR remain unclear. This study aimed to analyze long-term outcomes following TAVR using meta-analysis.

**Methods:** A literature search was performed with MEDLINE, EMBASE, Cochrane Library, Web of Science, and Google Scholar through December 2022; studies reporting clinical outcomes of TAVR with follow-up periods of ≥ 8 years were included. The outcomes of interest were overall survival and/or freedom from structural valve deterioration (SVD). Surgical risk was assessed with the Society of Thoracic Surgeons (STS) predicted risk of mortality (PROM) score. A subgroup analysis was conducted for intermediate-/high-surgical risk patients only.

**Results:** Eleven studies including 5,458 patients were identified and analyzed. The mean age was 82.0 ± 6.5 years, and mean STS PROM score ranged from 2.9 to 10.6%. Survival rate at 5 and 10 years was 47.7% ± 1.4% and 12.1 ± 2.0%. Five studies including 1,509 patients were analyzed for SVD. Freedom from SVD at 5 and 8 years was 95.5 ± 0.7% and 84.9 ± 3.1%. Similar results for survival and SVD were noted in the subgroup analysis of intermediate-/high-risk patients.

**Conclusions:** Following TAVR, approximately 88% of patients died within 10 years, whereas 85% were free from SVD at 8 years. These date suggest that baseline patient demographic have the greatest impact on survival, and SVD does not seem to have a prognostic impact in this population. Further investigations on longer-term outcomes of younger and lower-risk patients are warranted.

**What is new?:**

- Meta-analysis of time-to-event data was performed to assess long-term outcomes following transcatheter aortic valve replacement in a large population.
- Six studies, including 4,910 patients with follow-up periods of more than 20 years were identified and analyzed.
- Nearly 88% of patients who underwent transcatheter aortic valve replacement died within 10 years, while 85% remained free from structural valve deterioration at 8 years.

**Perspective Statement What are the clinical implications?:**

- This meta-analysis demonstrated excellent freedom from structural valve deterioration in intermediate- to high elderly risk patients.
- Baseline patient characteristics correlated with high mortality at 10 years.
- Further investigation into the outcomes in younger and lower-risk populations is imperative to evaluate the future expanded indications for transcatheter aortic valve replacement.

## Introduction

Transcatheter aortic valve replacement (TAVR) has been established as an alternative to surgical aortic valve replacement (SAVR) for most patients with severe aortic stenosis. Current practice guidelines recognize TAVR as a safe and effective option for patients across the surgical risk spectrum, for patients aged ≥65 years in the United States and ≥75 years in Europe[1, 2]. Although several randomized controlled trials have demonstrated short- and mid-term non-inferiority between the treatment with TAVR and SAVR in intermediate and low-surgical risk patients [3-5], data on long-term outcomes are still limited. Therefore, long-term durability of TAVs remains largely uncertain and there are concerns regarding the lack of data on long-term mortality and structural valve deterioration (SVD). In this context, we conducted a systematic review and meta-analysis to evaluate the long-term results beyond five years following TAVR.

## Methods

This study complies with the Preferred Reporting Items for System and Reviews and Meta-Analysis statement standards[6]. Studies were included if the following criteria were met;

1. The study design was randomized or nonrandomized, retrospective or prospective observational study with matched or unmatched populations;
2. The study included patients with aortic stenosis who underwent TAVR;
3. The study reported survival and/or incidence of SVD or freedom from SVD;
4. Kaplan-Meier curves were available with numbers at risk present ≥ eight years.

The included studies were not restricted to any language. The eligible studies were identified using a 2-level strategy. First, databases including MEDLINE, EMBASE, Cochrane Library, Web of Science, and Google Scholar were searched through November 2022 using Web-based search engines. Search terms included “transcatheter aortic valve replacement”, “transcatheter aortic valve implantation”, “transcatheter heart valve (THV)”, and “transcatheter aortic valve”. Second, relevant articles were manually identified through secondary sources, including the first identified studies’ reference lists. Two authors (ST and JS) independently reviewed extracted publications. Studies were included only if two investigators agreed. Two independent investigators (ST and JS) evaluated the risk of bias in individual studies using the Joanna Briggs Institute (JBI) critical appraisal checklist (Supplemental Table 1). The following information was extracted: authors, year of publication, year of enrollment/procedure performed, sample size, age, follow-up periods, predicted risk of mortality by the Society of Thoracic Surgeons (STS) predictaed risk of mortality (PROM) score[7], New York Heart Association class III/IV heart failure and left ventricular ejection fraction at the time of TAVR, history of diabetes mellitus, hypertension, peripheral vascular disease, chronic obstructive pulmonary disease, renal failure, myocardial infarction, atrial fibrillation, and stroke, previous coronary artery bypass graft or any cardiac surgery and pacemaker implantation, type of implanted valve and approach, 30-day mortality, cardiovascular mortality, and incidence of SVD.

**Table 1.**
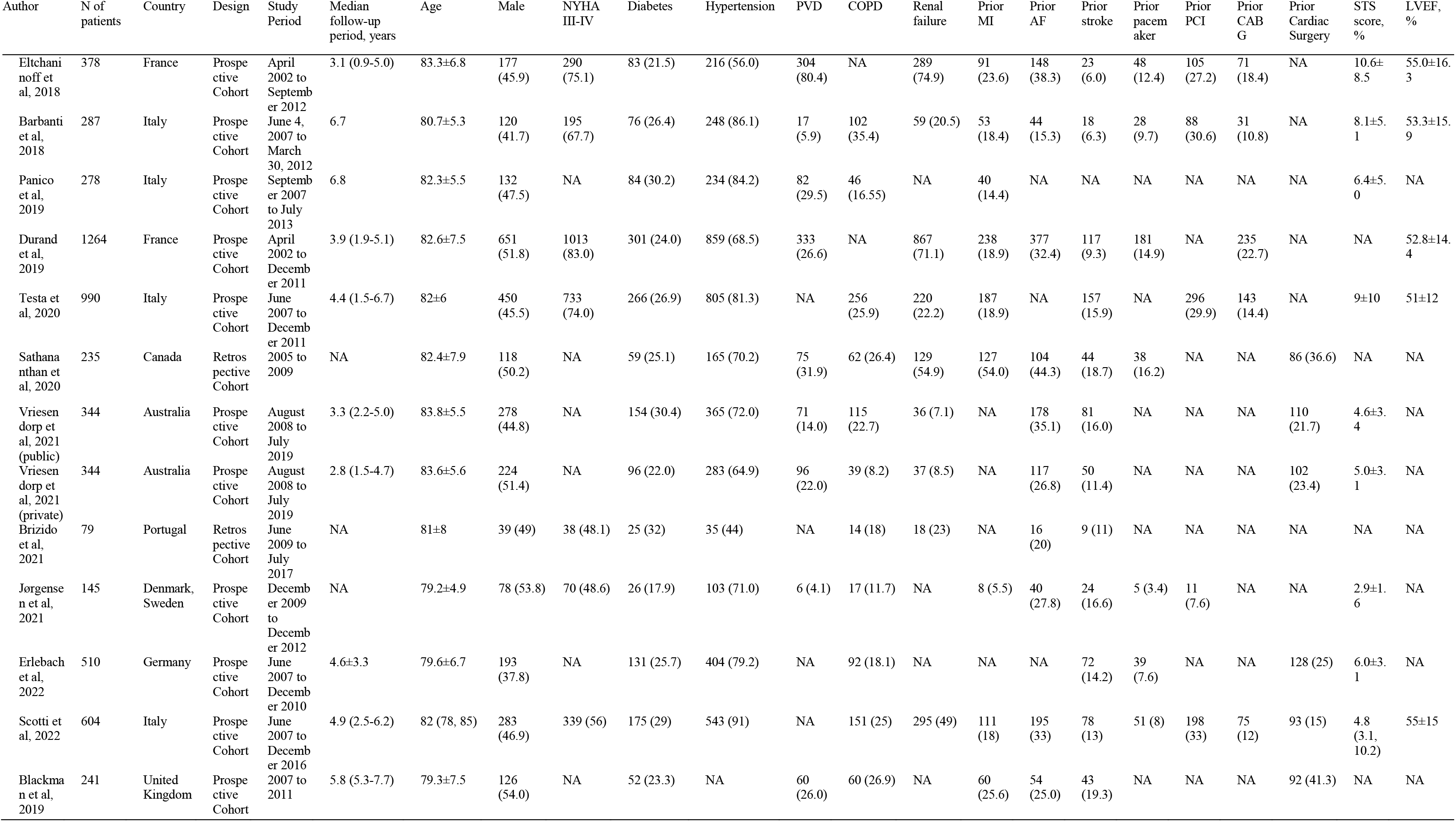
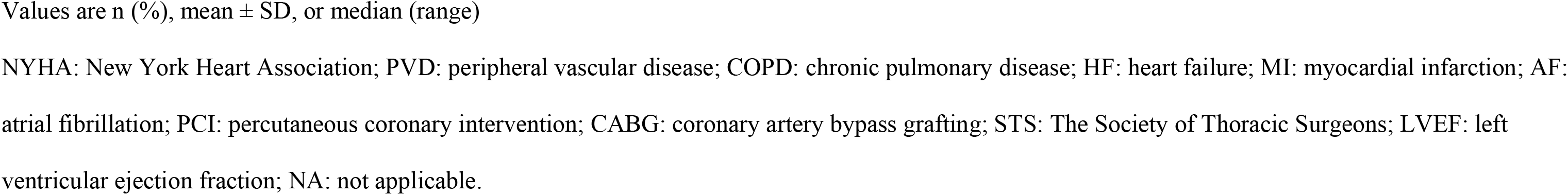
Summary of included studies and patient characteristics.

### Outcomes of interest

The primary outcome of this study was overall survival. The secondary outcome of interest was freedom from SVD.

### Definition of SVD

The European guidelines task force committee definition was used by all studies[8]. Moderate SVD was present when any of the following was present; 1) mean trans-prosthetic gradient ≥ 20 mmHg and < 40 mmHg; 2) ≥ 10 mmHg and < 20 mmHg increase from baseline; 3) moderate new or worsening intra-prosthetic aortic regurgitation. Severe SVD was present when any of the following was present; 1) mean trans-prosthetic gradient ≥ 40 mmHg; 2) ≥ 20 mmHg increase from baseline; 3) severe new or worsening intra-prosthetic aortic regurgitation.

### Statistical analysis

We analyzed data from the included studies based on the approach demonstrated by Liu *et al*. to obtain a merged Kaplan-Meier curve from individual Kaplan-Meier curves[9]. First, raw data coordinates (time, survival probability, or freedom from SVD) were extracted from Kaplan-Meier curves. If the incidence of moderate and severe SVD was reported separately, only moderate SVD data was included to avoid underestimating the incidence of SVD given the low incidence of severe SVD [10, 11]. Second, individual patient data in each study were reconstructed from data coordinates and the numbers at risk at given timepoint using the R package “IPDfromKM”. Finally, we merged individual patient data of all eligible studies, and drew the Kaplan-Meier curve. The analysis was conducted with R Statistical Software (version 4.2.1, Foundation for Statistical Computing, Vienna, Austria). Median values in each study were converted to mean using the quantile estimation method reported by McGrath et al. [12]. The converted mean data were combined into one group by Cochrane’s formula (https://training.cochrane.org/handbook/current/chapter-06). In addition, we also performed a subgroup analysis of patients at intermediate- or high-surgical risk.

## Results

### Literature selection results

A total of 11,987 studies were identified, published from 2002 to 2016, after removal of duplicates and evaluation based on the selection criteria, 12 studies were included in the meta-analysis (Figure 1). Eleven studies with a total of 5,458 patients were included in the survival analysis[11, 13-22]. One study was a randomized controlled trial and remaining ten studies were observational. Five studies with 1,509 patients were included in the analysis of freedom from SVD [10, 11, 16, 17, 19]. One study was a randomized controlled trial, and remaining studies were observational. Four of these five studies were also included in the survival analysis[11, 16, 17, 19]. The included studies are summarized in Table 1. The studies were conducted between 2002 and 2016. Only one study (the NOTION trial) included 145 patients with low-surgical risk (2.7% for survival analysis and 9.6% for SVD analysis of the study population).

**Figure 1.**
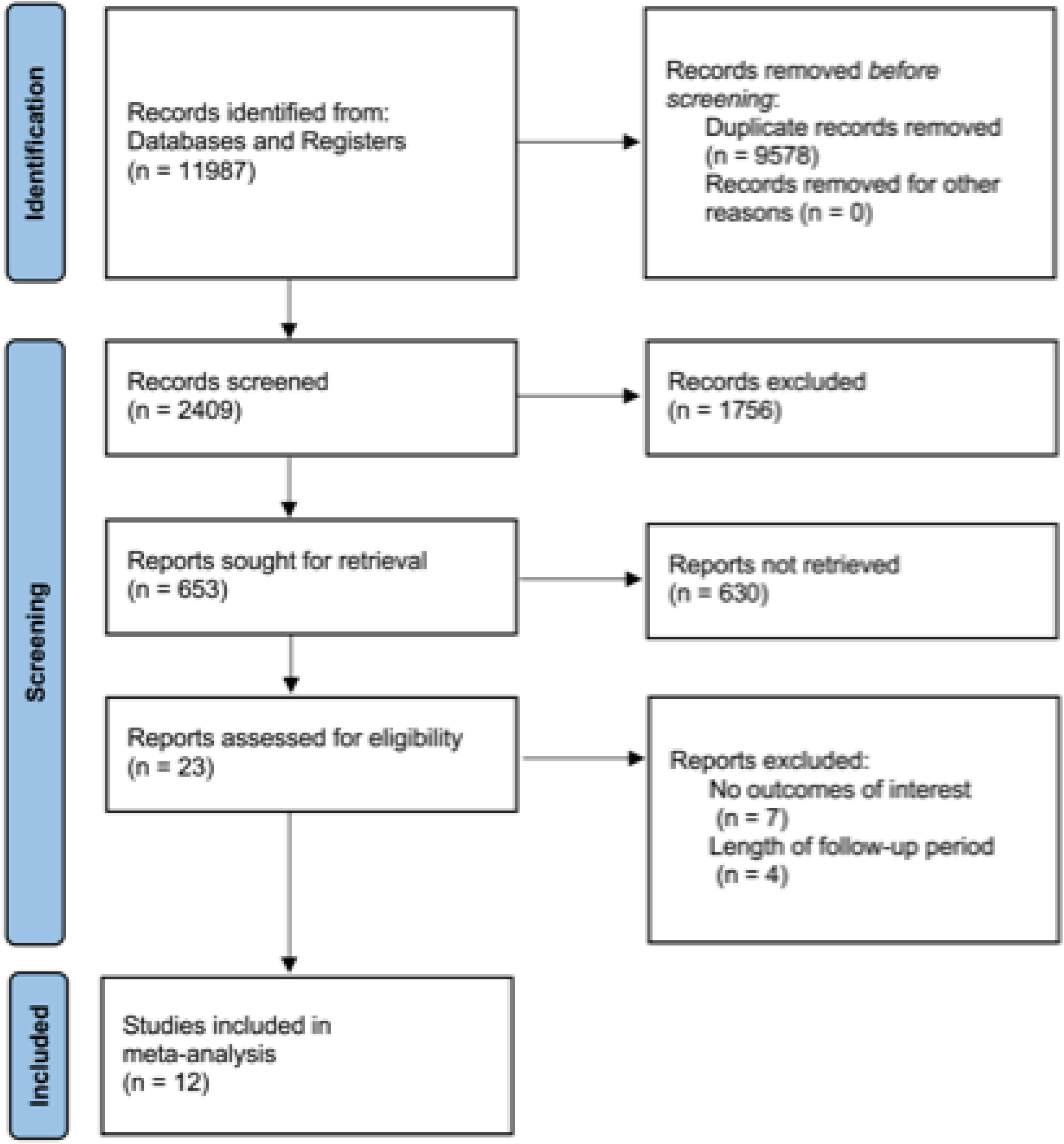
Preferred Reporting Items for Systematic Reviews and Meta-Analyses (PRISMA) flow diagram.

### Patient and procedure characteristics

Patient characteristics are summarized in Table 1. In the survival analysis, the longest follow-up ranged from 8 to 12.4 years among included studies, with a weighted median of 4.9 years. Pooled mean age was 82.0 ± 6.5 years, and the mean STS PROM score ranged from 2.9 to 10.6%. Regarding the implantation approach, 76.5% (4,175/5,458) of cases were transfemoral TAVR. In the freedom from SVD analysis, the mean age was 80.1±7.1 years, and a weighted median follow-up period was 5.8 years. The longest follow-up period ranged from 8 to 11 years. Valve types are described in Table 2. In five studies reporting freedom from SVD, 722 (47.8%) TAVRs were performed using SAPIEN^®^, SAPIEN XT^®^, or SAPIEN 3^®^ valves (Edwards Lifesciences, Irvine, California, USA), and 643 (42.6%) using CoreValve^®^ (Medtronic, Minneapolis, Minnesota, USA). Early-generation THVs (SAPIEN, SAPIEN XT, CoreValve) accounted for 90.5% of patients.

**Table 2.**
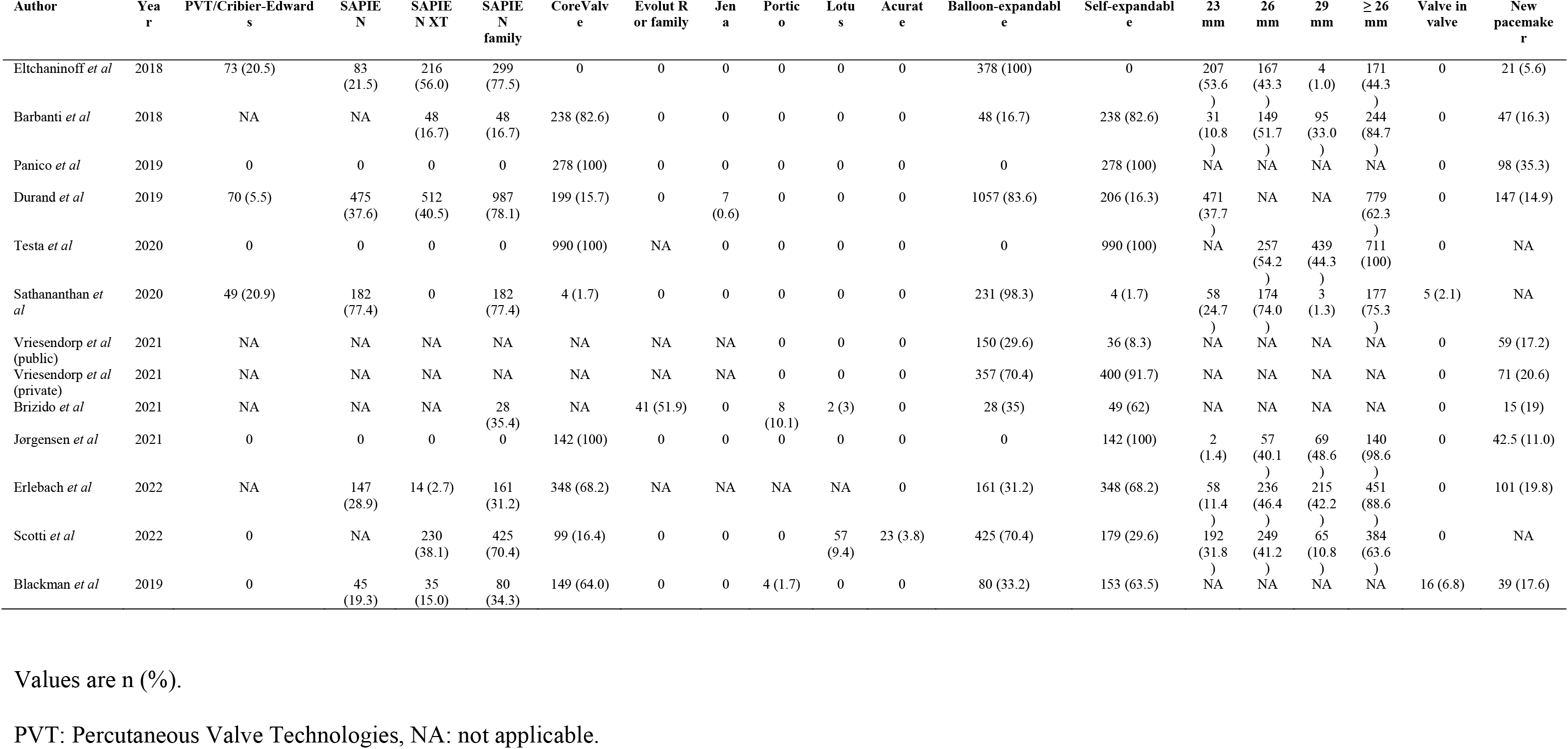
Procedural data.

### All-cause mortality and freedom from SVD

Survival rate and freedom from SVD in each study are summarized in Supplemental Tables 2A and 2B. Aggregated survival rate was 47.7 ± 1.4% and 12.1 ± 2.0% at 5 and 10 years, respectively (Figure 2A). Incidence rate of death was 13.8 per 100 person-years (i.e., 13.8% annually). Aggregated freedom from SVD was 95.5 ± 0.7% and 84.9 ± 3.1% at 5 and 8 years, respectively (Figure 3A). The 11 studies in the survival analysis did not provide information on the specific causes of death; however, 6 of the 11 studies disclosed the percentages of cardiovascular death to overall death, which ranged from 7.6% to 60.5%. Timing of death, the incidence of early and late mortality, and incidence of cardiovascular death are summarized in Supplemental Table 3.

**Figure 2.**
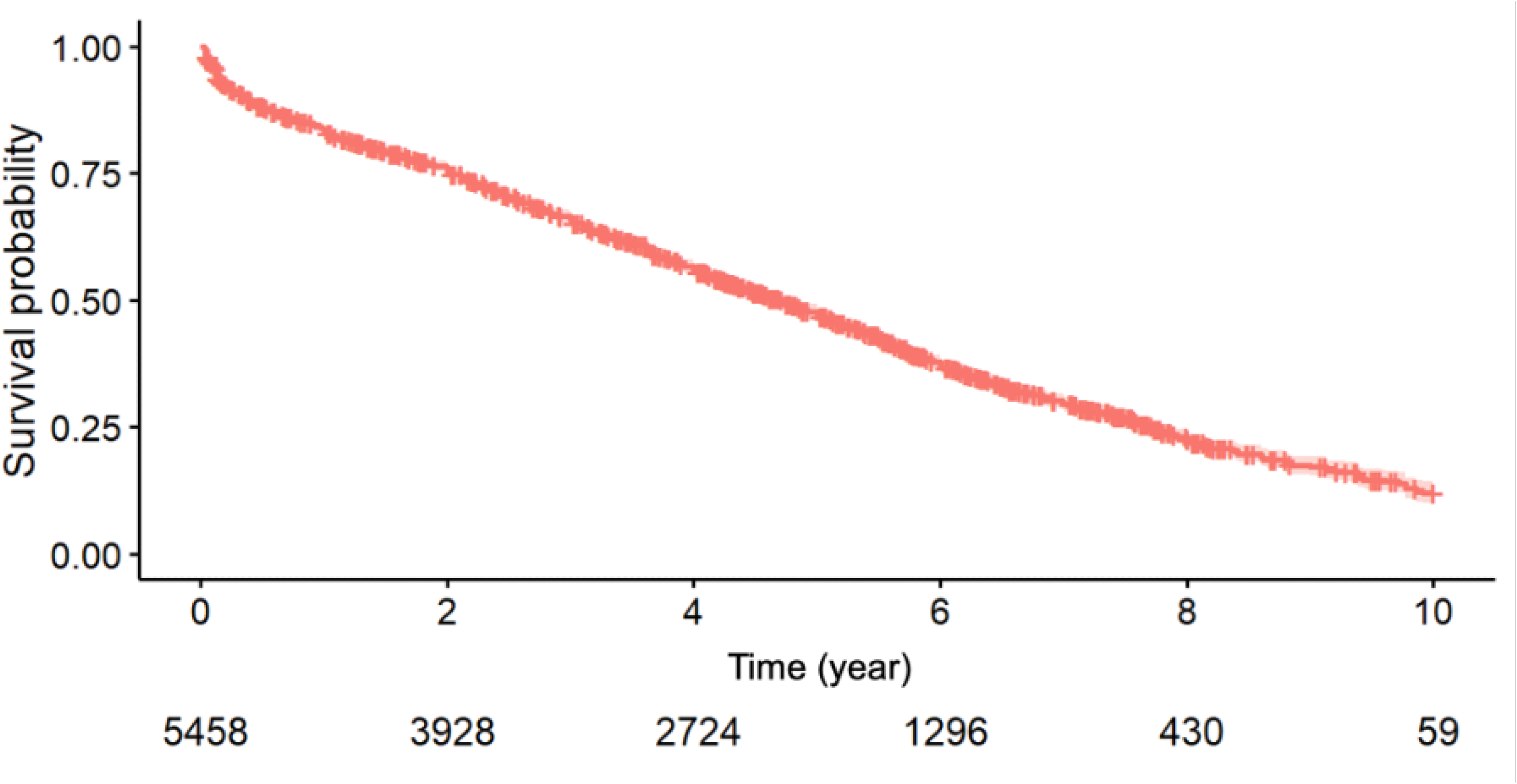
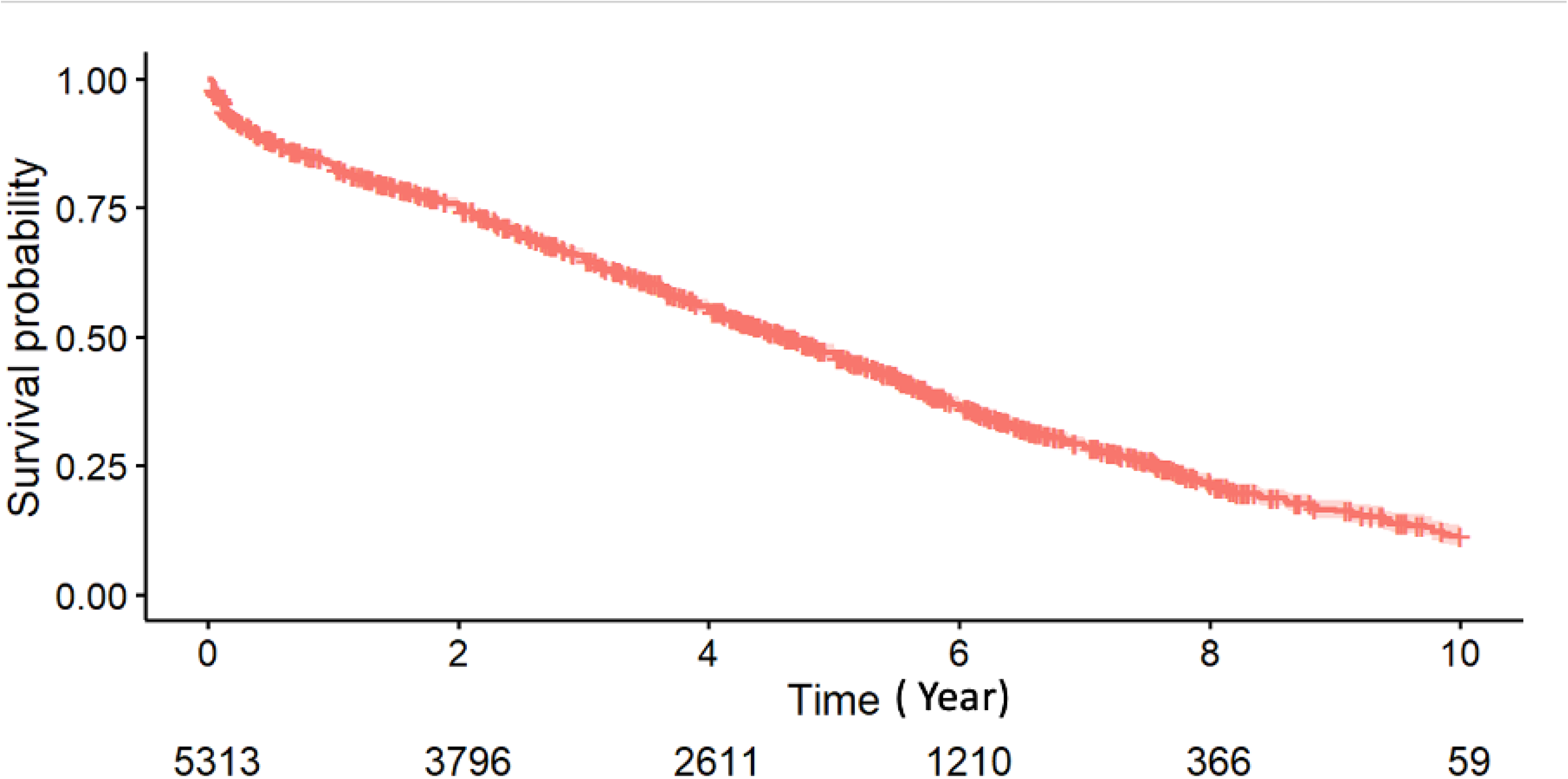
Kaplan-Meir curve describing overall survival following TAVR with 95% confidence intervals reconstructed from pooled individual patient data. TAVR: transcatheter aortic valve replacement. A: overall cohort B: cohort excluding patients with low surgical risk

**Figure 3.**
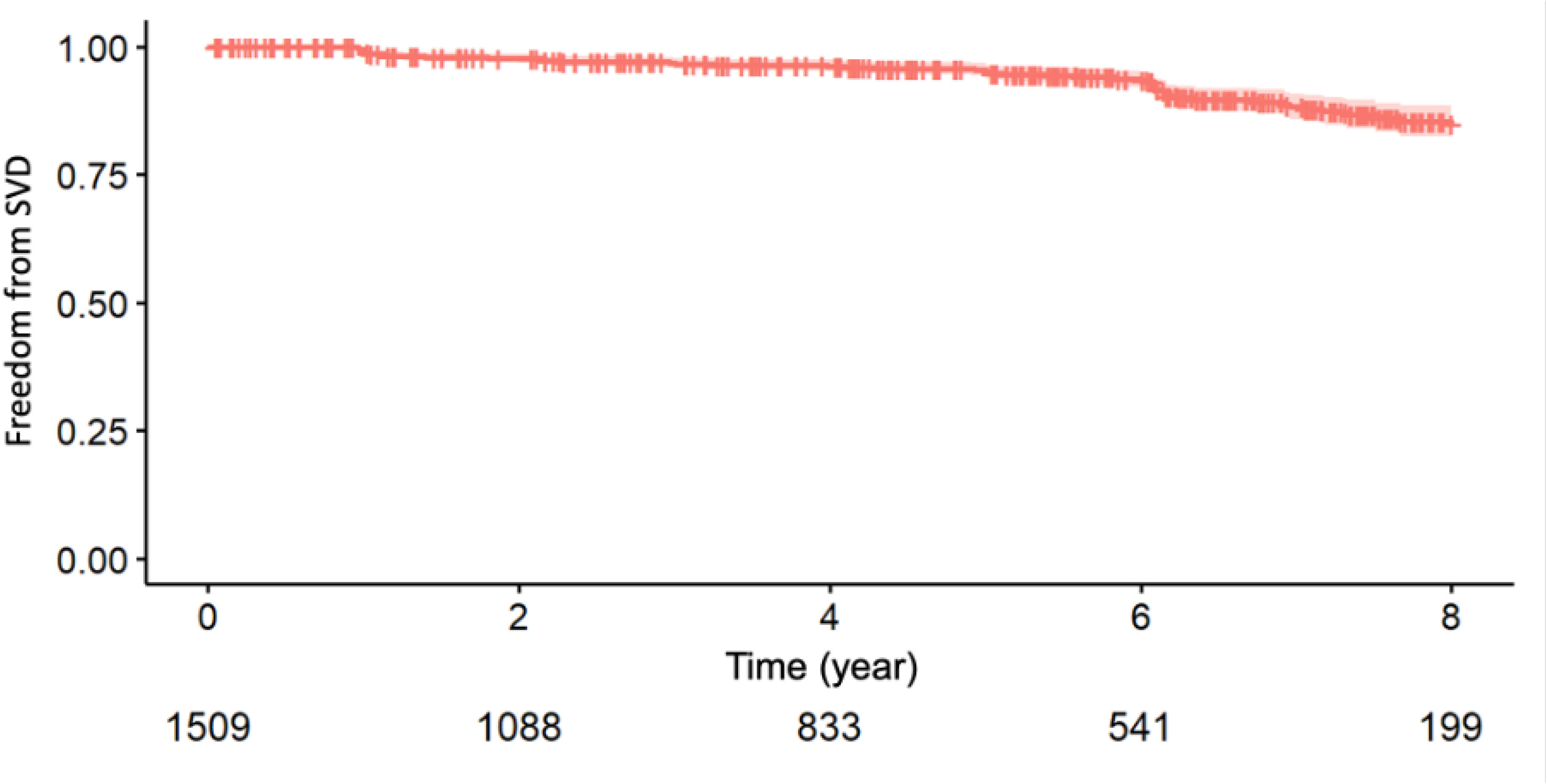
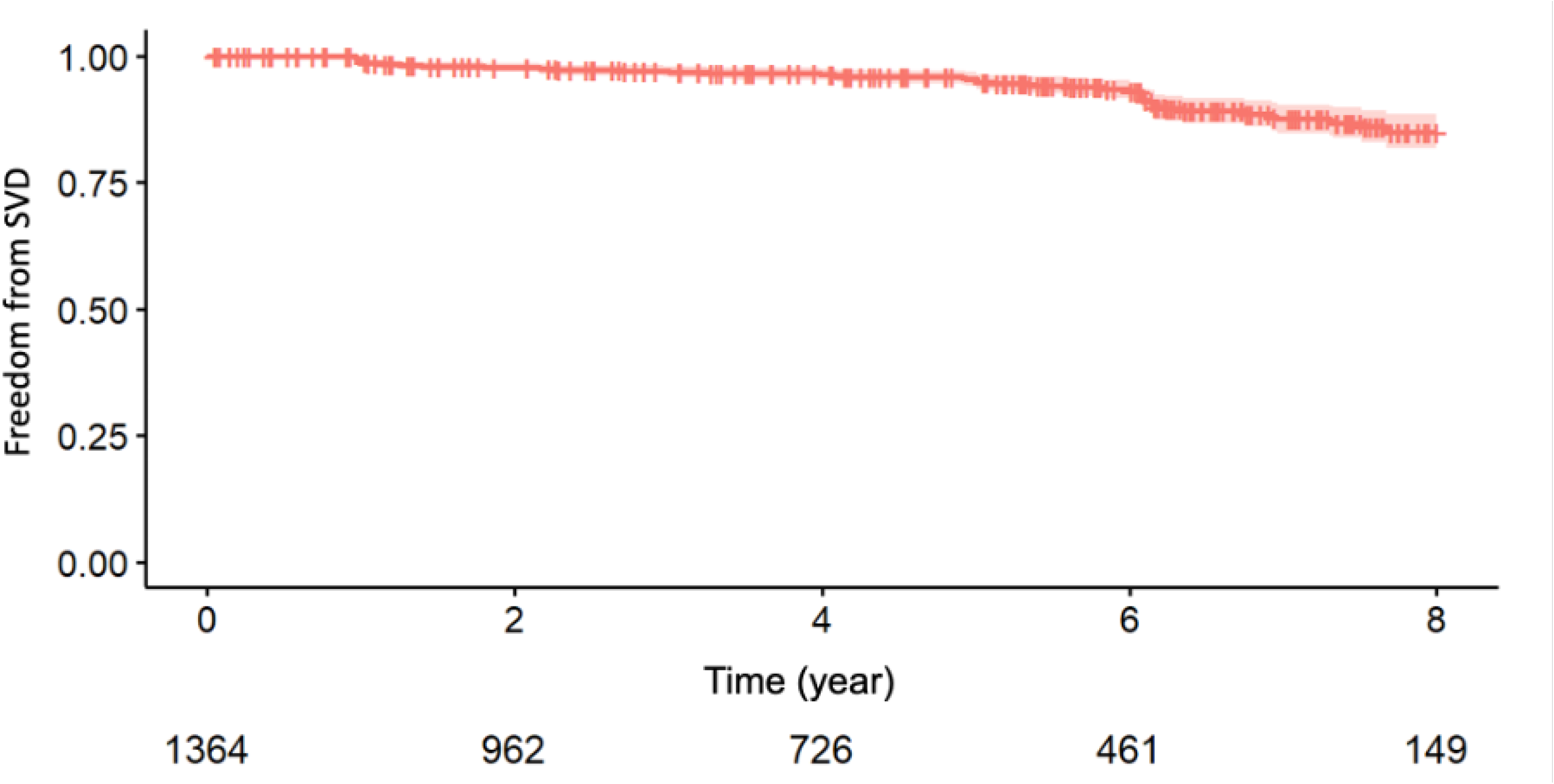
Kaplan-Meir curve describing freedom from SVD following TAVR with 95% confidence intervals reconstructed from pooled individual patient data. SVD: structural valve deterioration; TAVR: transcatheter aortic valve replacement. A: overall cohort B: cohort excluding patients with low surgical risk

### Subgroup analysis of intermediate- or high-surgical risk patients

In the subgroup analysis excluding the one study with low-surgical risk patients, the mean age was 82.1 ± 6.6 years old, and the mean STS PROM score ranged from 4.6 to 10.6%. Survival rate at 5 and 10 years was 47.0% ± 1.3% and 11.4 ± 2.2%, respectively (Figure 2B). Freedom from SVD at 5 and 8 years was 95.5 ± 0.7% and 85.0 ± 3.3%, respectively (Figure 3B).

## Discussion

In this meta-analysis of 11 studies (5,458 patients with an average age of 82 years and mostly high/intermediate-surgical risk), we report the following important findings: (1) overall survival following TAVR at 10 years was 12.1 ± 2.0%; (2) freedom from SVD at 8 years was 84.9 ± 3.1%.

### Long-term survival

The high late mortality rate in our study was mainly attributed to the elderly population with a short life expectancy and significant comorbidities reflected in the high STS PROM scores. This data must be interpreted based on the natural history of severe aortic stenosis, life expectancy of the matched population, and mortality following surgical aortic valve replacement (SAVR). If severe aortic stenosis is left untreated, the annual mortality rate of patients with untreated severe aortic stenosis is known to be approximately 25%[23]. Also, patients who did not undergo TAVR had 5-year mortality of 93.6%, which is higher than 10-year mortality among patients who underwent TAVR in this study[24].

Regarding long-term survival following SAVR, Malvindi et al. analyzed 1,870 patients aged 80 years and over with logistic EuroSCORE of 13.3 % who underwent SAVR and reported that the survival rate was 31% at 10 years, which was similar to the expected survival of a sex- and aged -matched population[25]. Similarly, in a study by Brennan et al. analyzing over 43,000 patients who underwent SAVR with either mechanical or bioprosthetic prosthesis primarily for aortic stenosis, the survival rate for high-risk patients was approximately 20% at 8 years[26]. In contrast, the NOTION trial included patients of similar age (mean age, 79) but low surgical risk and treated with self-expanding CoreValve, and the survival rate was higher with 51.8% at 8 years following TAVR[17]. Given the small number of patients in NOTION in our study, the survival rate was similar with or without low surgical-risk patients included, resulting in mostly intermediate-high risk patients with a higher mortality rate. Nonetheless, to understand the accurate survival of elderly patients undergoing TAVR with low-/intermediate-risk, especially over 75 years of age, a further analysis based on the surgical risk stratification is warranted. Moreover, since the cut-off age for TAVR is still controversial and there exists a difference between the two major guidelines[1, 2], understanding the long-term survival data in younger patients, such as those between 65-75 years of age, is essential before expanding the indication for TAVR.

Prosthesis-patient mismatch (PPM) is another important consideration. Despite technological advances in valve design, PPM is an unsolved complication following SAVR and TAVR. Although the impact of post-SAVR PPM on survival has been well-recognized, the evidence regarding post-TAVR PPM remains limited[27] with some data that PPM may influence the survival following TAVR[28]. The incidence of PPM appears higher with balloon-expandable valve than self-expandable valve, whereas the impact on survival seems to be higher with self-expandable valve than balloon-expandable valves[29, 30]. Further assessment of PPM incidence and its prognostic significance stratified by valve type are needed.

### Freedom from SVD

Our meta-analysis of SVD at 8-year provides insights into long-term THV performance. Studies reporting durability of the THVs according to the new standardized criteria of the 2017 EAPCI/ESC/EACTS consensus, moderate SVD was reported at 3.6-14.9%, severe at 0.8-3.7% at follow-up of 5-8 years[10, 11, 16, 17, 19].

Due to the extremely low late survival and the resultant influence on the SVD rate as competing events, our results should be interpreted with caution. Nevertheless, we demonstrate acceptable durability of TAVR at 8 years with over 85% of patients were free from SVD. We elected to use the 2017 consensus document definition of SVD as it better corresponded with the era of the trials included in this meta-analysis. The Valve Academic Research Consortium 3 definitions were published subsequently and may provide a more consistent definition of SVD going forward[31].

A main clinical concern currently is whether TAVR valves show non-inferior durability compared to bioprosthetic SAVR valves. The frequency of SVD based on reintervention of the Carpentier-Edward PERIMOUNT aortic valve at 10 and 20 years were 1.9% and 15% overall[32] in a study of 12,569 patients (mean age,71). Few randomized trials have compared long-term durability of TAVR valves vs bioprosthetic SAVR and results are conflicting. In NOTION trial, 280 patients (mean age, 79.1) with low surgical risk were randomized. At 8 years of follow-up SVD was lower in TAVR than SAVR (13.9% vs 28.3%). In the post hoc analysis of pooled date from the CoreValve US High Risk Pivotal and SURTAVI trials, a total of 2,099 patients (mean age, 82.1) were included. At 5 years of follow-up, cumulative incidence of SVD treating death as a competing risk was lower in patients undergoing TAVR than SAVR (TAVR, 2.2%; SAVR 4.38%). SVD was associated with a two-fold increased risk of 5-year all-cause mortality[32]. Ueyama et al. performed a network meta-analysis to compare the durability of the THVs (balloon-expandable and self-expandable) and SAVR prosthesis[33]. They reported that self-expandable valves demonstrated significantly larger effective orifice area, lower mean aortic valve gradient, and lower incidence of SVD compared with balloon-expandable and SAVR valves at 5 years.

Both TAVR valves suffered an increased risk of aortic regurgitation and re-intervention compared with SAVR valves. The latest generation bioprosthesis for SAVR, the INSPIRIS RESILIA^®^ aortic valve (Edwards Lifesciences LLC, Irvine, CA) using calcification-prevent technology, has demonstrated 100% of freedom from SVD in 689 patients at 5 years, expecting further longer-term durability[34]. Similarly, SAPIEN 3 Ultra RESILLIA which has been recently launced has enhanced calcium blocking properties and future THVs also may be able to provide better durability with technological advances. For the accurate understanding of the THV durability, even longer follow-up, assessment stratified by valve type and generation, and echocardiographic data are essential.

In our study, both balloon-expandable valve and self-expandable valves were used (only balloon-expandable in two, only self-expandable in one, and both valves in two studies). A comparative study between CoreValve and SAPIEN up to 7 years was performed by Deutsch et al., demonstrated favorable outcomes in SVD with CoreValve[35]. (11.8% in CoreValve vs 22.6% in SAPIEN). This result may be explained by supra-annular configuration of self-expandable valves, which provides a larger effective orifice area than balloon-expandable valves with intra-annular leaflet design. Devices of different generations were also included in our study. Early-generation THVs (SAPIEN, SAPIEN XT, CoreValve) accounted for 90.5% of patients in the current SVD analysis. Further study is warranted regarding the incidence of SVD in patients treated with newer generation valves and performance of each THVs should be assessed individually.

### Limitations

This study has several limitations. First, our study included only one randomized study and has the inherent limitations of meta-analysis related to the retrospective observational studies. Therefore, we could only perform a single arm analysis of patients underwent TAVR. Second, follow-up on valve durability was performed in a survival cohort with death exerting a competing risk and it may have biased and underestimated the SVD rate. Third, as in all studies with long-term follow-up, patient cohort and utilized THVs (type or generation) and approaches differ from the current population and technologies. Causes of death were not available in most of the studies, resulting in the difficulty to assess the impact of cardiovascular events on mortality.

## Conclusions

Among elderly TAVR recipients in the intermediate- to high-risk category, approximately 88% died within 10 years, which is markedly superior compared to the untreated severe AS population. Over 85% of patients were free from SVD at eight years, suggesting acceptable durability of the early generation TAVR valves. Further longer-term investigations in younger and lower-risk patients with newer generation TAVR valves are warranted.

## Data Availability

All data generated or analysed during this study are included in this published article and supplementary information files.

## Glossary of Abbreviations

SAVR: surgical aortic valve replacement
STS: the Society of Thoracic Surgeons
SVD: structural valve deterioration
TAVR: transcatheter aortic valve replacement
THV: transcatheter heart valve
PROM: predictaed risk of mortality PPM prosthesis patient mismatch

